# Comparing existing and novel methods for estimating etiology-specific diarrheal disease incidence in hybrid surveillance studies

**DOI:** 10.1101/2025.11.11.25339698

**Authors:** Maria Garcia Quesada, Alexander Breskin, James A. Platts-Mills, David Benkeser, Patricia B. Pavlinac, Sean R. Galagan, Lance A. Waller, Benjamin A. Lopman, Elizabeth T. Rogawski McQuade

## Abstract

Accurate estimates of infectious disease incidence are critical for designing studies of public health interventions, including vaccines. Hybrid surveillance studies estimate incidence by enrolling cases in medical facilities, estimating population denominators in the community, and adjusting for healthcare seeking behaviors, which is necessary to minimize bias. The Enterics for Global Health (EFGH) Study aimed to generate updated incidence estimates of *Shigella* diarrhea among children in preparation for vaccine trials. We conducted a simulation to evaluate approaches for healthcare seeking adjustment and uncertainty estimation in hybrid studies and applied these methods to EFGH. Adjusting for healthcare seeking using the inverse of individual-level propensity scores for healthcare seeking greatly reduced bias compared to the inverse of the marginal probability for healthcare seeking. M-estimation and bootstrap 95% confidence intervals both had at least nominal coverage of the truth across scenarios. Monte Carlo 95% simulation intervals had nominal coverage in some scenarios but not all. When applied to EFGH, M-estimation confidence intervals around fully adjusted incidence estimates were narrower than bootstrap. Computation time for M-estimation using the *geex* R package was significantly higher than bootstrap or Monte Carlo, making bootstrap an appealing option for valid results and ease of use.

## 1. Background

*Shigella* is a major cause of diarrhea morbidity and mortality, particularly among children in low resource settings, and multiple *Shigella* vaccine candidates are currently in the pipeline.^1–3^ Accurate estimates of *Shigella* disease incidence are needed to appropriately design the upcoming vaccine trials, but cohort studies, in which incidence can be directly estimated, are time and cost prohibitive.^4–6^ Hybrid surveillance designs are an appealing alternative in which cases are enrolled in medical facilities, population denominators are estimated in the community, and incidence is then estimated using some adjustment for healthcare seeking behaviors.^7^ Adjusting for care seeking is complicated because care seeking is driven by several factors, including disease severity, sociodemographic factors, as well as access to, and quality of, care. Access to care differs across settings and by study design: in rigorous individually randomized clinical trials, care seeking will likely be facilitated and incentivized, whereas in passive surveillance studies, care seeking patterns likely reflect those that occur naturally in the community. Therefore, the healthcare seeking adjustment in hybrid surveillance designs is a critical source of potential bias when estimating adjusted incidence rates.

Hybrid surveillance studies have commonly been used to generate incidence estimates of typhoid fever to inform typhoid conjugate vaccine and antibiotic use.^8–10^ The specific methods used by each study varies, but the core methodology involves upweighting the number of cases captured in medical facilities to account for those that did not seek care. Most simply, this has been done using the inverse of the proportion of children in the community reporting seeking healthcare for an equivalent syndrome. A similar approach was used in two studies of diarrhea etiology among children in low resource settings, the Global Enteric Multicenter Study (GEMS) and the Vaccine Impact on Diarrhea in Africa (VIDA) studies.^11,12^ To estimate uncertainty around their estimates, typhoid studies have generally relied on a Monte Carlo approach, using simulations to sample from a distribution of each adjustment factor, including the healthcare seeking proportion.^8,9^ We identify two opportunities to improve on these methods, in the healthcare utilization adjustment and in the estimation of uncertainty around the adjusted incidence rate.

First, healthcare utilization adjustments applied as simple proportions within one or two stratifying features can fail to capture the full heterogeneity in healthcare seeking behaviors by disease severity and sociodemographic characteristics.^13^ Given the potential impact of small differences in such proportions on adjusted incidence rates, proper adjustment is critical. Instead of stratifying by a large number of factors, many of which do not neatly fall into discrete categories (i.e., disease severity), we propose using a regression to model individual-level propensity-for-healthcare-seeking scores. These propensity scores quantify how likely each case enrolled in a study facility was to seek care based on a set of sociodemographic and clinical characteristics. Then, we can use the inverse of these scores to upweight cases and account for those with similar characteristics who did not present to care.^14^

Second, the methods used to estimate uncertainty (e.g., 95% confidence intervals) in prior hybrid studies have not been validated.^8–10^ Because parameters from multiple datasets are combined in the incidence estimate, usual generalized linear model-based standard error estimates are not available. Monte Carlo simulations, as previously applied in the typhoid studies, are based on unverifiable assumptions about the distributions of adjustment factors. Bootstrapping may be valid but has not been empirically validated for the type of complex survey designs used to adjudicate population-based incidence. Wald-type bootstrap confidence intervals require the assumption that the parameter estimator is normally distributed, which may only be valid when sample size is large.^15^ Percentile bootstrap confidence intervals enjoy some robustness to departures from normality, but also require large sample size for validity and may still demonstrate poor coverage in finite samples.^16^ An alternative method for estimating robust standard errors is M-estimation. M-estimators are solutions to a set of estimating equations.^17^ In this case, an M-estimator can be used to “fuse” the different datasets collected in hybrid studies (i.e., facility case data, population enumeration data, healthcare utilization data) via one system of equations. Through these equations, it can adjust for sampling of the numerator and denominator, adjust for the probability of healthcare seeking by using a propensity score model, and estimate incidence, all while appropriately accounting for all sources of uncertainty.^18–20^ Here, we evaluated methods for healthcare seeking adjustment and uncertainty estimation in hybrid studies, with the aim to identify methods that minimize bias and improve precision of infectious disease incidence estimates. Specifically, we aimed to estimate incidence of *Shigella* diarrhea in the Enterics for Global Health (EFGH) *Shigella* surveillance study, as results will be used by investigators to inform upcoming *Shigella* vaccine trial design.^21,22^ To evaluate the proposed methods, we also conducted a simulation to mimic the data structure in EFGH and hybrid studies generally. First, we evaluated the performance of propensity score weights to adjust for healthcare seeking compared to weighting using the marginal probability. Then, we developed a data fusion design with M-estimation to estimate confidence intervals around incidence estimates and compared that to those estimated with Monte Carlo simulations and bootstrapping.

## 2. Methods

### 2.1 Enterics for Global Health (EFGH) Shigella Surveillance Study Data

EFGH is a hybrid surveillance study that aimed to generate incidence estimates of *Shigella* diarrhea among children in seven country sites in preparation for *Shigella* vaccines approaching Phase 2b/3 trials.^21^ Broadly, it includes active facility-based surveillance, population enumeration of the facility catchment areas, and a healthcare utilization survey (HUS) conducted among a subset of the those enumerated.

Data collection occurred between 2022 and 2024. Country sites included Bangladesh, Kenya, Malawi, Mali, Pakistan, Peru, and The Gambia, and study facilities captured a total catchment area of approximately 2.8 million persons, ranging from ∼100,000 persons in Blantyre, Malawi and Iquitos, Peru to 1.6 million persons in Karachi, Pakistan. Children aged 6-35 months of age presenting with diarrhea at one of the study health facilities and who resided within the pre-defined study area were eligible to be enrolled as cases.^23^ Diarrhea was defined as ≥3 abnormally loose or watery stools within 24 hours. Screening and enrollment questionnaires captured clinical and sociodemographic information from each eligible child. Stool samples were then collected from those enrolled (prior to receiving antibiotic therapy, if applicable) and etiology was attributed to 16 enteropathogens by qPCR. *Shigella* was also isolated via culture and serotyped by PCR.

Population enumeration surveys were conducted in the catchment area of each study facility, as defined by geographic or administrative boundaries.^24^ To do so, grid maps used by WorldPop (an open access spatial demographic dataset) were first overlaid on the study catchment area for each site, and square grids were aggregated into clusters such that the average total population per cluster was approximately 500. Clusters were then randomly selected for enumeration. Each household within each selected cluster was visited at least once, and those that consented were asked about the number and age of each child under five years of age living in the house.

If a visited household had at least one child in the study target age range of 6-35 months who had diarrhea in the past two weeks, that household was invited to participate in the HUS.^24^ The HUS captured if and where those children sought care, as well as similar diarrhea symptoms and sociodemographic information as that captured among the facility cases.

### 2.2 Simulated Hybrid Study Data

To validate the performance of propensity score weights to adjust for healthcare seeking behaviors, as well as each uncertainty estimation method, we simulated data for a hybrid study parametrized based on the observed values in EFGH. The population in the catchment area and diarrhea cases occurring over the course of a two-year study were simulated separately, and all parameters for the simulation are shown in Supplementary Table 1 and Supplementary Table 2.

The population at risk was simulated by first simulating clusters within the catchment area. Each cluster was assigned an income level (low, middle, high) and a number of households within the cluster. Then, each household was assigned a wealth quintile, conditional on income level of the cluster, and a number of children <5 years of age. Each child was assigned an age in months, whether they experienced diarrhea in the two weeks prior, and if so, symptoms associated with the episode including number of loose stools on worst day of diarrhea, any fever, any vomiting, and blood in stool. A healthcare seeking probability was assigned to each child reporting diarrhea using a logistic regression model predicting the probability of healthcare seeking based on age, symptoms, and wealth quintile. As a proxy for distance to healthcare facilities, cluster-level variability was incorporated by adding random normal noise to probabilities at the cluster level. As a proxy for other unmeasured factors, individual-level variability was incorporated using random normal noise. Those determined to have sought care were further randomly assigned to seeking care at a study facility or other facility. A random subset of clusters was selected for enumeration by the study and a random subset of households in enumerated clusters were selected for being reached by the study team.

Simulated diarrhea cases occurring over the course of a two-year study were each assigned an age in months and a wealth quintile. Using the same strategy as for diarrhea cases in the simulated population at risk, each case was assigned symptoms associated with the episode, a healthcare seeking probability with individual-level variability, and if they sought care, whether they did so at a study facility. Lastly, diarrhea etiology due to *Shigella* was randomly assigned using a logistic regression model predicting the probability of *Shigella* etiology in a diarrhea case based on age, symptoms, and wealth quintile. If cases sought care at a study facility, they were randomly selected to being enrolled in the study.

The main simulation scenario, referred to as the average scenario, was based on average parameters across all EFGH country sites. Additionally, we simulated relative high- and low-healthcare seeking scenarios, which used all the same parameters except the logistic regression model for predicting healthcare seeking was based on the EFGH sites with highest and lowest healthcare seeking, respectively. We also simulated high- and low-case scenarios, where the true number of diarrhea cases were adjusted such that resulting *Shigella* incidence rates were higher than the highest incidence EFGH site and lower than the lowest incidence EFGH site, respectively. We generated 1,000 simulations for each of the five scenarios. The target of inference for each simulated data set was the true incidence rate of *Shigella* in the simulated population, which varied slightly across simulations, as would be expected across sites in a multisite study.

### 2.3 Healthcare seeking adjustment

A propensity score model was used to estimate the probability of accessing care given a set of symptoms and sociodemographic characteristics. The inverse of these estimates was then used to upweight the facility cases to account for those with the same profile that did not access care (see Appendix 2-1 for basic illustration of the method).

The propensity score model was a logistic regression model with coefficients based on data from the HUS. The outcome was defined as caregiver report of seeking care for diarrhea at a healthcare facility (inpatient or outpatient facility within the catchment area). In the simulated data, predictors included age, wealth quintile, number of loose stools on worst day of diarrhea, any fever, any vomiting, and blood in stool. In the EFGH data, predictors included all variables with a hypothesized association with healthcare seeking behaviors that were captured by both the facility enrollment survey and the HUS: age in months, sex, country site, wealth quintile, number of loose stools on worst day, number of days with fever, number of days with vomiting, number of times vomited on worst day, and caregiver report of blood in stool. See Appendix 2-2 for more detail on EFGH variable and interaction selection.

We estimated the bias in incidence adjusted for healthcare seeking by calculating the average relative difference between the estimated incidence rate and the truth in each simulation. We compared the bias in each scenario when using propensity score weights to weighting using the marginal probability of healthcare seeking. To evaluate the impact of extreme weights, we also compared the bias to using propensity score weights truncated at the 99^th^ and 95^th^ percentiles (i.e., set all weights above the 99^th^ and 95^th^ percentiles to the 99^th^ and 95^th^ percentile weight values, respectively). The least biased approach was used in subsequent simulations to evaluate uncertainty estimation methods.

We hypothesized the healthcare seeking adjustment would be less important for more severe definitions of diarrhea, which would likely be the primary endpoint in a vaccine trial, as opposed to an endpoint of any diarrhea. To evaluate this hypothesis, we estimated the percent increase in incidence estimates in EFGH after adjusting for healthcare seeking for all diarrhea and for moderate-to-severe diarrhea (MSD) as defined in the GEMS study (i.e., diarrhea with dysentery, dehydration, or hospitalization).^11^

### 2.4 Adjusted incidence and confidence intervals

To estimate adjusted incidence and confidence intervals, we developed a fusion design M-estimator and compared the M-estimator confidence intervals to those generated by bootstrap and Monte Carlo simulation. M-estimators enable the estimation and combination of statistical parameters from different data sources using stacked estimating functions and allow a closed-form standard error estimate.^18,19^ Here, we integrated the three components of a hybrid surveillance study – facility-based case surveillance, population enumeration data, and healthcare utilization data – to estimate adjusted incidence.

### 2.4.1 Adjustments for cases (numerator)

Cases were defined as children in the target age range with *Shigella*-attributable diarrhea detected by qPCR, based on a pre-specified pathogen quantity cutoff.^25^ To estimate the true number of cases from the observed (i.e., enrolled) number of cases, we upweighted cases to account for both healthcare seeking behaviors and facility sampling.

The overall healthcare seeking weight for each case *c* was the product of the weight to account for healthcare seeking at any facility (*w*_*HC seek,c*_) and a weight to account for the proportion that sought care at a study facility enrolling cases among those that sought care at any facility (*w*_*HC seek at study facility,c*_). The weight to account for any healthcare seeking was the inverse of their propensity score, from the propensity score model described above. The weight to account for healthcare seeking at a study facility was the inverse of the site-stratified proportion of children who sought care at a study facility out of all those who sought care.

The facility sampling weight for each case was the product of a screening weight (*w*_*screen,c*_) and an enrollment weight (*w*_*enroll,c*_). These weights aim to capture the proportion of cases who would have been eligible to participate in the study but did not due to lack of consent, no longer being at the facility, or other administrative reasons (i.e., presented overnight or during a weekend when EFGH screening did not occur, or presented after the site-specific monthly enrollment cap had been met). The screening weight was the inverse of the proportion of cases who were eligible for screening and were screened, stratified by site and facility. The enrollment weight was the inverse of the proportion of cases who met the enrollment criteria and were enrolled, stratified by site, facility, and type of diarrhea (i.e., dysentery or watery based on caregiver report).

#### 2.4.2 Adjustments for child-years at risk (denominator)

Child-years at risk were estimated separately for each facility, then summed for each country site. First, the adjusted number of children 6-35 months at risk were estimated by multiplying the number of children enumerated by 1) the inverse of the proportion of households enumerated in each randomly selected cluster *q* (*w*_*HH enum,q*_), and 2) the inverse of the proportion of the total study area that was enumerated (*w*_*area enum*_), based on geography. Then, the adjusted number of children 6-35 months was multiplied by the number of days of clinical surveillance by the facility and divided by the number of days in a year.

#### 2.4.3 Adjusted incidence

Assembling all the adjustment factors, the adjusted numerator *N* was estimated using weights for each case *c* and facility *h*, and the adjusted denominator *D* was estimated using weights for each catchment area cluster *q. N* and *D* were then used to estimate the adjusted *Shigella* incidence *I* as follows:

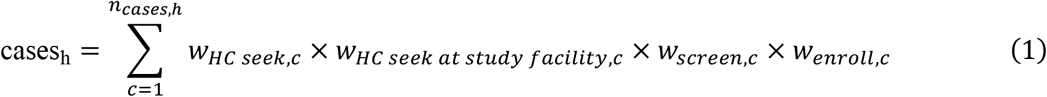

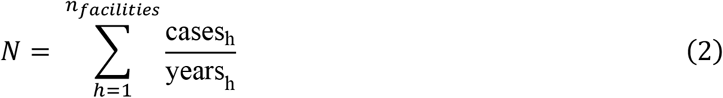

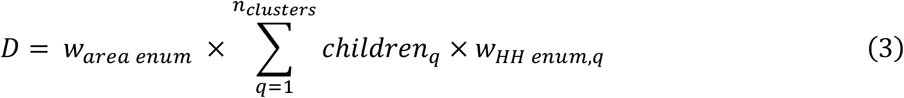

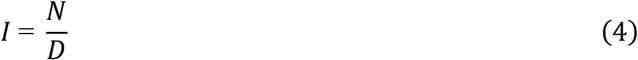

Here, *children*_*q*_ is the number of children enumerated in each cluster selected for enumeration, cases_h_ is the adjusted number of cases captured by each facility, and years_h_ is the number of years each facility recruited cases.

#### 2.4.4 M-estimation

In this section, we describe how we can adapt the above calculation for adjusted incidence to be computed by M-estimation software. The basic structure of an M-estimator 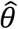 is as the solution in *θ* to an equation:^26^

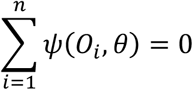

where *ψ* is a *k*-dimensional estimating function, *O*_*i*_ is the observed data for each unit *i, θ* is a *k*-dimensional parameter, and *n* is the total number of data units (i.e., rows). To solve for *k* parameter(s) in the estimating function(s), these must be arranged into *k* equations such that one side of each equation is set to zero.

One set of unknown parameters we need to solve for are the coefficients of the propensity score model, which is a logistic regression model generally expressed as:

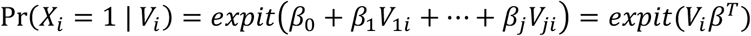

Where *X*_*i*_ is an indicator of healthcare seeking, *V*_*i*_ = (1, *V*_1*i*_, …, *V*_*ji*_) is a row-vector of all predictors included in the model, and *β* = (*β*_0_, *β*_1_, …, *β*_*j*_) is a row-vector of their respective coefficients. To include this in an M-estimator and solve for *j* coefficients *β*, we consider the following system of *j* + 1 score equations denoted by *ψ*_1_:^26^

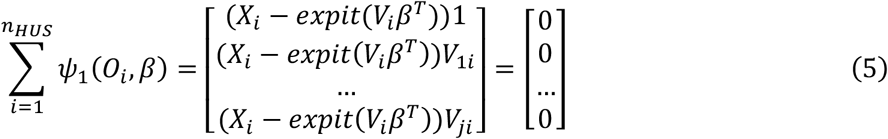

The solution for *β* in the M-estimator above is the same as the solution for the logistic regression model if solved using maximum likelihood estimation. These coefficients, estimated using the HUS data (which has *n*_*HUS*_ rows), can then be used to estimate *w*_*HC seek,c*_ among facility cases.

We similarly use logistic regression models to estimate *w*_*HC seek at study facility,c*_ from the HUS data with estimating equations *ψ*_2_:

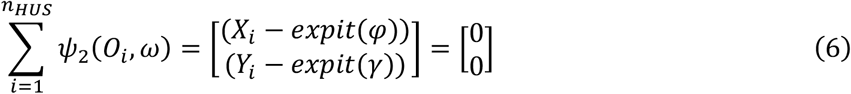

Where *X*_*i*_ is again healthcare seeking at any facility, *Y*_*i*_ is healthcare seeking at a study facility, and *ω* = (*φ, γ*) (i.e., the intercepts for each model). We can estimate the marginal weight for seeking care at any facility as 1/*expit*(*φ*) and the marginal weight for seeking care at a study facility as 1/*expit*(*γ*). The estimated weight for seeking care at a study facility can be divided by the weight for seeking care at any facility, yielding a weight for seeking care at a study facility among those who seek care.

For *w*_*area enum*_, we use estimating equation *ψ*_3_ and the population enumeration data, which has *n*_*pop*_ rows. Here, *Z*_*i*_ is whether a cluster was enumerated and *δ* is the model intercept:

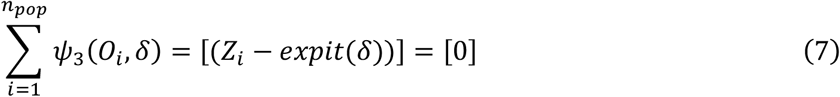

For the simulation, models in *ψ*_2_ and *ψ*_3_ are intercept only models (as shown here), and for EFGH these include only covariates for country site to estimate site-specific weights.

The remaining weights needed to estimate adjusted incidence (i.e., screening weight for cases [*w*_*screen,c*_], enrollment weight for cases [*w*_*enroll, c*_], and weight for percent of households enumerated per population cluster [*w*_*HH enum,c*_]) are known values, rather than estimated from a sample and do not contribute to the overall uncertainty.

To allow M-estimation software to incorporate uncertainty from each unknown adjustment parameter into the resulting adjusted incidence, all parameters should be estimated within a single M-estimator. Therefore, the three datasets (i.e., facility case data, population enumeration data, healthcare utilization data) need to be stacked into a single dataset for analysis by M-estimation software, such that the total number of rows *n* corresponds to the sum of the number of rows across the three datasets (i.e., *n* = *n*_*HUS*_ + *n*_*pop*_ + *n*_*case*_). The stacked dataset includes all relevant variables from the three datasets, and indicator variables to differentiate the rows of the stacked dataset that correspond to each of the three original datasets. This data structure is illustrated in Table 1.

**Table 1.** Illustration of data structure required for M-estimation.

| *hus*<br>(indicator<br>variable<br>for HUS<br>data) | *case*<br>(indicator<br>variable<br>for<br>facility<br>case<br>data) | *pop*<br>(indicator<br>variable<br>for<br>population<br>enumeration<br>data) | *Sought*<br>*care for*<br>*diarrhea*<br>*a*<br>(0 =<br>no, 1 =<br>yes) | *Sought*<br>*care for*<br>*diarrhea*<br>*a* at<br>*study*<br>*facility*<br>(0 =<br>no, 1 =<br>yes) | *Age*<br>(months<br>) | *Wealt*<br>*h*<br>*quintil*<br>*e* | *Vomitin*<br>*g*<br>(0 = no,<br>1 = yes) | *Enrolme*<br>*nt weight* | *Number of*<br>*children*<br>*enumerate*<br>*d in*<br>*cluster* |
| --- | --- | --- | --- | --- | --- | --- | --- | --- | --- |
| 1 | 0 | 0 | 1 | 1 | 7 | 3 | 0 | 999 | 999 |
| 1 | 0 | 0 | 0 | 0 | 29 | 2 | 0 | 999 | 999 |
| 1 | 0 | 0 | 1 | 0 | 10 | 5 | 1 | 999 | 999 |
| 1 | 0 | 0 | 0 | 0 | 12 | 1 | 1 | 999 | 999 |
| 1 | 0 | 0 | 1 | 1 | 32 | 1 | 0 | 999 | 999 |
| 0 | 1 | 0 | 999 | 999 | 23 | 5 | 1 | 3.54 | 999 |
| 0 | 1 | 0 | 999 | 999 | 16 | 5 | 0 | 1.59 | 999 |
| 0 | 1 | 0 | 999 | 999 | 9 | 3 | 0 | 1.59 | 999 |
| 0 | 1 | 0 | 999 | 999 | 14 | 1 | 0 | 2.78 | 999 |
| 0 | 1 | 0 | 999 | 999 | 26 | 2 | 1 | 2.78 | 999 |
| 0 | 0 | 1 | 999 | 999 | 999 | 999 | 999 | 999 | 29 |
| 0 | 0 | 1 | 999 | 999 | 999 | 999 | 999 | 999 | 53 |
| 0 | 0 | 1 | 999 | 999 | 999 | 999 | 999 | 999 | 78 |
| 0 | 0 | 1 | 999 | 999 | 999 | 999 | 999 | 999 | 11 |
| 0 | 0 | 1 | 999 | 999 | 999 | 999 | 999 | 999 | 16 |

Not real data and not a comprehensive illustration of all variables included in analyses. 999 represents “not applicable.”

With our estimating equations for unknown parameters and our stacked dataset, we can then assemble our full M-estimator to compute adjusted incidence and confidence intervals. First, we include equations from (5), (6), and (7) to estimate the unknown adjustment parameters. Second, we include additional estimating equations that solve for the adjusted numerator *N*, denominator *D*, and incidence *I*, equivalent to equations (2), (3), and (4). By including these additional equations, M-estimation software effectively automates the delta method calculations needed to obtain standard error estimates that account for the uncertainty across all estimated parameters. Because we are now using a single stacked dataset rather than the individual datasets, we must use indicator variables to differentiate which parts of the dataset should be used for which estimating equations. For example, we multiply *ψ*_1_ by *hus*_*i*_, an indicator variable to indicate whether each row in the data is a HUS response, such that propensity score model coefficients are only estimated with the HUS data in our stacked dataset. Then, using indicator variable *case*_*i*_, we use the resulting coefficients to generate propensity scores among cases; the inverse of these scores corresponds to *w*_*HC seek,i*_. Similarly, we multiply *ψ*_2_ by *hus*_*i*_ and *ψ*_3_ by *pop*_*i*_. Our resulting M-estimator solves for *θ* = (*β, ω, δ, N, D, I*).

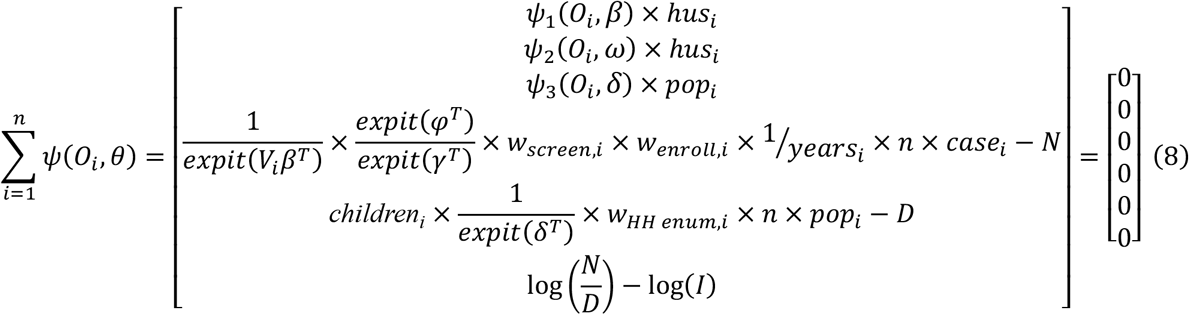

Here, *years*_*i*_ is the surveillance time for the facility that enrolled each case and *children*_*i*_ is the number of children enumerated in each cluster. The estimating equations are evaluated over all rows in the data, but the denominator of the resulting estimator depends on the estimating equation itself. Therefore, to return the correct numerator (*N*) and denominator (*D*) we multiply the corresponding equations by the number of rows (*n*) in the data.

To estimate robust confidence intervals around the parameters of interest, the M-estimator uses a sandwich variance estimator 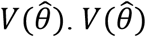 is defined as:^26^

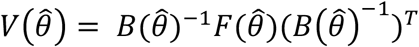

Where 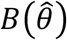 is a vector of estimated means of the partial derivatives (the Jacobian) of the estimating functions, and 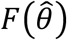 is the estimated mean of the outer product of the estimating functions:

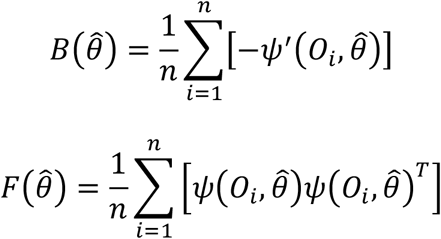

The analysis was done in R version 4.4.1 using the *geex* package, which is designed for M-estimation and requires a specific format of data and valid estimating functions to output the resulting parameters and their sandwich variance.^27^ The *geex* package includes a parameter that allowed the M-estimator to account for clustering in the numerator due to repeat cases being enrolled, and in the HUS due to these being completed in different geographic clusters.

#### 2.4.5 Bootstrap

We also estimated incidence and confidence intervals using the bootstrap, with stratified resampling by EFGH country (or simulation) and dataset (cases vs population enumeration/HUS). To account for clustering among cases, the unit of resampling was child given that in EFGH about 10% of enrolled cases were repeat enrollments. To account for clustering in the population enumeration and HUS responses, the unit of resampling was geographic cluster. Within each bootstrap resample, we computed all adjustment factors with uncertainty (i.e., *w*_*HC seek,c*_, *w*_*HC seek at study facility,c*_, and *w*_*area enum*_) and calculated incidence. Adjustment factors from known values (i.e., *w*_*screen,c*_, *w*_*enroll,c*_, and *w*_*HH enum,q*_) were kept fixed across resamples. Using 1,000 bootstrap resamples, we estimated both percentile (95%) and Wald style confidence intervals around the resulting incidence estimates.

#### 2.4.6 Monte Carlo simulations

We also estimated incidence and simulation intervals using Monte Carlo simulations to emulate the methodology used in prior hybrid studies of Typhoid incidence.^8,9,13^ This approach aims to introduce uncertainty into adjusted incidence estimates by sampling from a range of values for each adjustment factor with uncertainty (i.e., *w*_*HC seek,c*_, *w*_*HC seek at study facility,c*_, and *w*_*area enum*_). Given that these weights are each the inverse of a proportion bound by 0 and 1, we sampled the proportions from a logit-normal distribution. These were parametrized by the mean and prediction standard errors of each proportion in the data. For *w*_*HC seek,c*_, these parameters were case-specific based on the propensity score estimated by the healthcare seeking model. For *w*_*HC seek at study facility,c*_ and *w*_*area enum*_, these parameters were country site specific. After estimating total adjusted case counts by multiplying enrolled cases by their corresponding weights, we sampled the adjusted counts from a Poisson distribution to account for variability in the cases. We generated 10,000 simulations and from those estimated 95% simulation intervals based on the distribution of resulting incidence estimates.

#### 2.4.7 Comparison of uncertainty estimation methods

In each scenario of the simulated data, we estimated four different incidence estimates each with incremental adjustments. First, we estimated the incidence of cases enrolled at study facilities, where the only adjustments were applied to the denominator. Second, we estimated incidence of all cases who sought care at study facilities, adjusting for screening and enrollment. Third, we estimated incidence of cases who sought care, adjusting for care seeking at study facilities among those who sought care. And fourth, we estimated total incidence, adjusting for healthcare seeking at any facility. For each, we estimated 1) the coverage of each uncertainty estimation method by quantifying the percent of simulations where 95% confidence intervals (95% CI), or simulation intervals for Monte Carlo, capture the truth, and 2) the precision of each method by comparing the median and interquartile range (IQR) of confidence interval widths. Lastly, we applied all methods to the EFGH data and compared precision and computation time across methods.

All analyses were performed on a MacBook Pro (2023) with an Apple M3 Pro chip, 12 CPU cores and 18 GPU cores, 36 GB of unified memory, and a 1 TB SSD storage. The operating system was macOS Sequoia 15.3. Monte Carlo simulations and bootstrap leveraged parallelization over 11 cores, while M-estimation must be run in a single process.

## 3. Results

### 3.1 Simulation results

#### 3.1.1 Healthcare seeking adjustment

Adjusting using the marginal probability for healthcare seeking resulted in the most precise estimates across scenarios but was also the most biased (Table 2). In the high and low care seeking scenarios, the truth (15.77 cases per 100 person-years on average) was overestimated by 3.69% (95% CI: 3.35, 4.04) and 9.51% (95% CI: 8.84, 10.19), respectively (Table 2). Conversely, the truth was underestimated by -2.17% (95% CI: -2.60, -1.73) in the average scenario (15.78 cases per 100 person-years on average), -2.32% (95% CI: -2.72, -1.92) in the high case scenario (47.30 cases per 100 person-years on average), and -2.57% (95% CI: -3.13, - 2.02) in the low case scenario (3.61 cases per 100 person-years on average; Table 2).

**Table 2.**
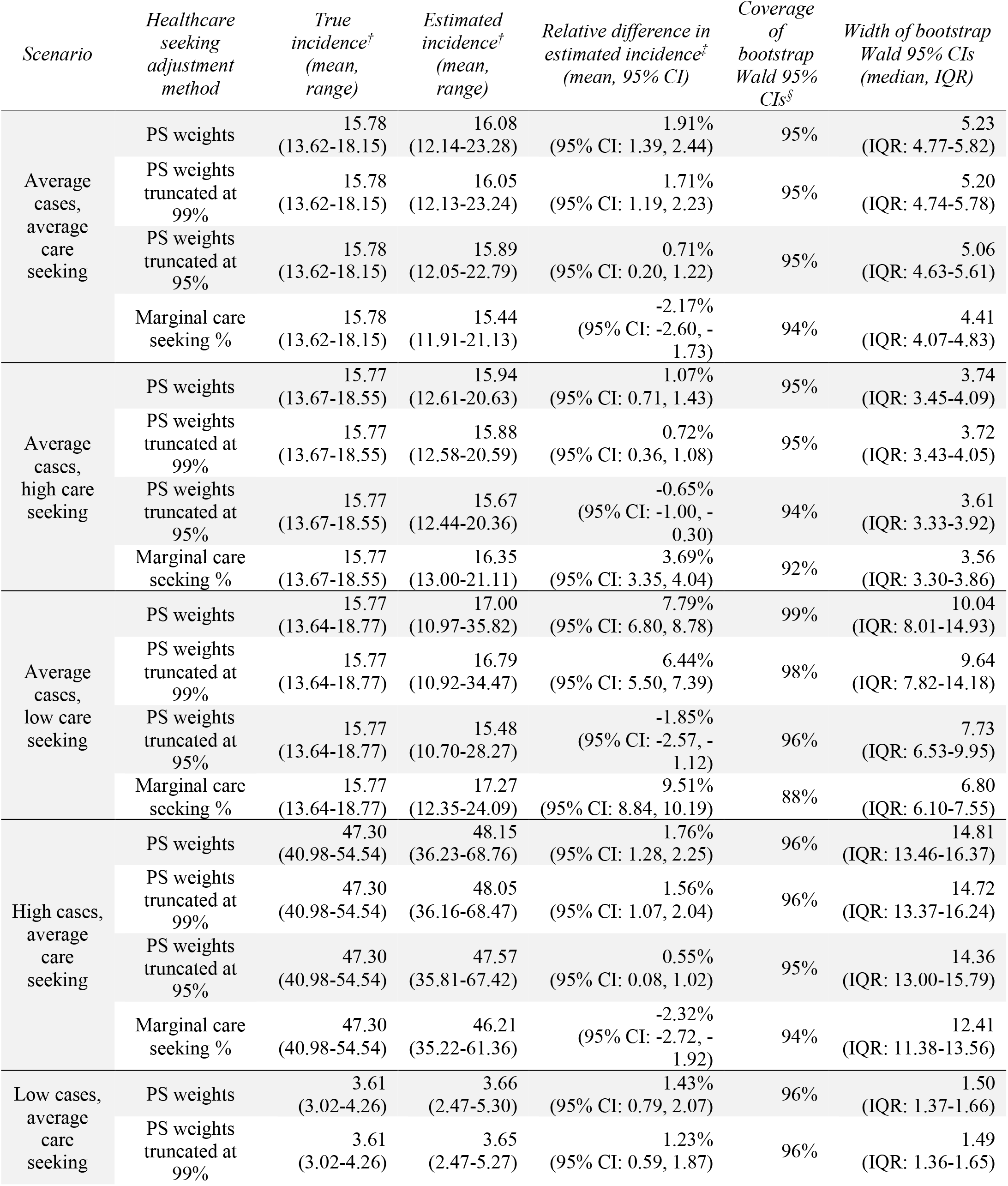
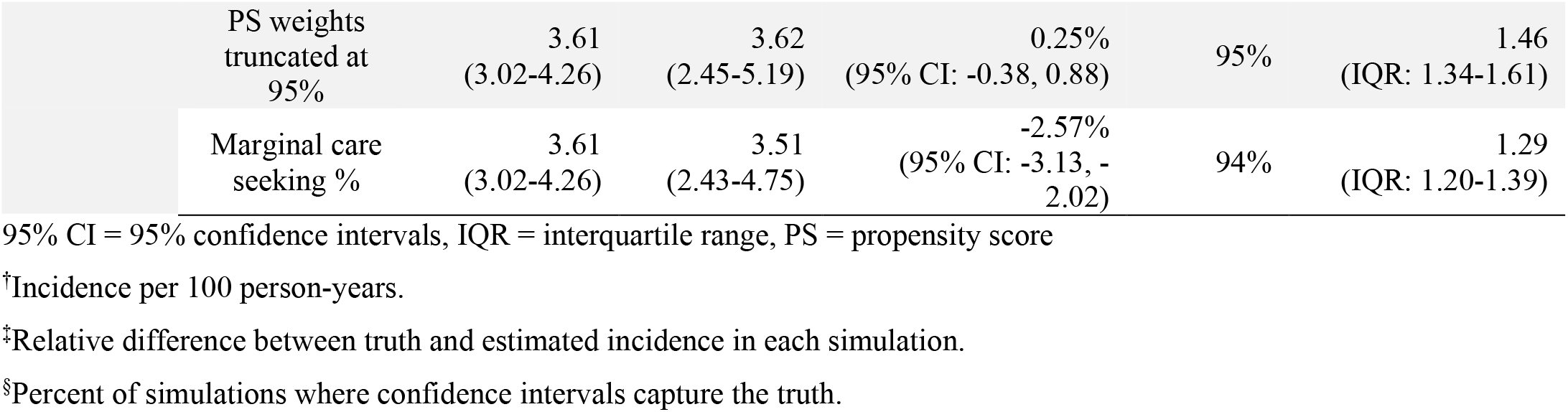
Simulation results comparing bias and precision of estimated total incidence using different approaches to adjust for healthcare seeking.

Using propensity score weights resulted in less bias than the marginal probability, but incidence was still overestimated on average (Table 2). Truncating weights at the 99^th^ percentile slightly reduced this bias, but truncating at the 95^th^ percentile performed best across scenarios; the relative difference between the estimated and true incidence per 100 person-years was 0.71% (95% CI: 0.20, 1.22) in the average scenario, -0.65% (95% CI: -1.00, -0.30) in the high care seeking scenario, -1.85% (95% CI: -2.57, -1.12) in the low care seeking scenario, 0.55% (95% CI: 0.08, 1.02) in the high case scenario, and 0.25% (95% CI: -0.38, 0.88) in the low case scenario (Table 2). Even though incidence was significantly over- or underestimated in certain scenarios when using truncated weights at the 95^th^ percentile, the mean bias was lower than with all other adjustment approaches.

Regardless of truncation, Wald-type bootstrap 95% confidence intervals around propensity score adjusted incidence had close to nominal coverage; the truth was within the bounds in 94-99% of simulations across scenarios, compared to 88-94% when using the marginal probability for healthcare seeking (Table 2). Truncating the weights improved precision, with propensity score weights truncated at 95^th^ percentile having the best precision compared to untruncated weights and similar coverage (Table 2).

Given the low bias, high coverage, and improved precision of using propensity score weights truncated at the 95^th^ percentile, these are used in the subsequent analyses of uncertainty estimation methods. Even with truncated weights, the adjustment for healthcare seeking is responsible for a large difference between incidence of cases who sought care and total incidence (in the average scenario, median 5.6 cases per 100 person-years [IQR: 5.3-5.9] to median 15.8 [IQR: 14.8-16.9]) (Figure 1).

**Figure 1.**
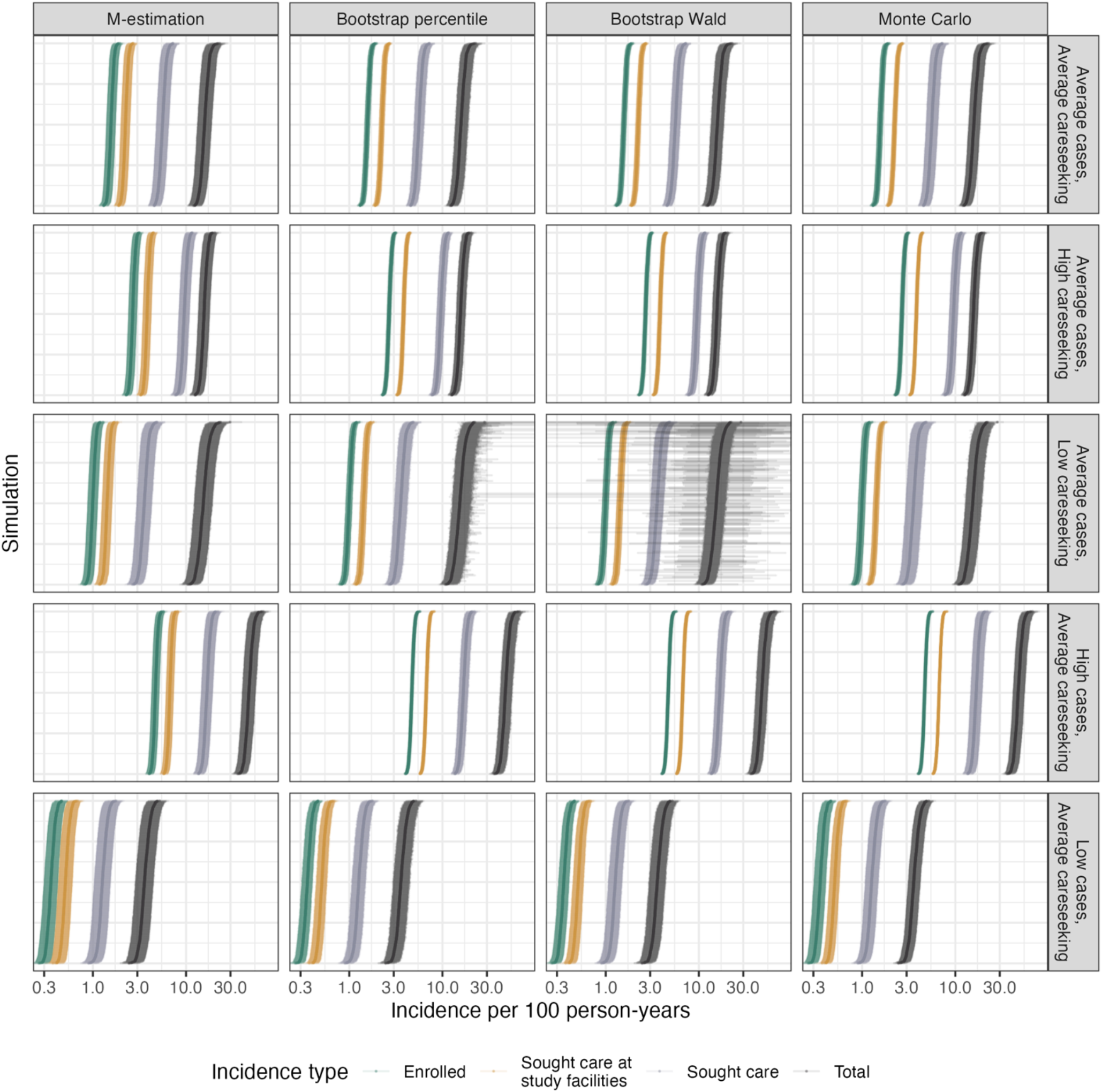
Estimated incidence rates, for each level of incidence adjustment, across simulations and scenarios. Points and shaded areas represent the estimated incidence rates and 95% confidence intervals with each uncertainty estimation method. X-axis is on the log scale.

#### 3.1.2 Adjusted incidence and uncertainty estimation

In the average scenario, M-estimation had above nominal coverage across the different incidence adjustment levels (97-100%) (Table 3). Bootstrap percentile and Wald-type confidence intervals had above nominal coverage for incidence of cases enrolled (100%) and cases who sought care at study facilities (100%), and close to nominal coverage for cases who sought care (95-96%) and total incidence (94-95%) (Table 3). Monte Carlo simulation intervals also had above nominal coverage for incidence of cases enrolled (100%), cases who sought care at study facilities (100%), and cases who sought care (99%), and slightly below nominal coverage for total incidence (93%) (Table 3). Results were generally consistent across scenarios with two exceptions. First, M-estimation had slightly below nominal coverage for total incidence in the low care seeking scenario (93%) (Table 3). Second, Monte Carlo simulation interval coverage for total incidence ranged from 95% in the high case scenario to 87% in the low case scenario (Table 3).

**Table 3.** Percent of confidence or simulation intervals that capture the truth across scenarios, for different incidence adjustment levels and uncertainty estimation methods.

| Scenario | Incidence adjustment level | M-estimation | Bootstrap percentile | Bootstrap Wald | Monte Carlo |
| --- | --- | --- | --- | --- | --- |
| Average cases, average care seeking | Enrolled | 100% | 100% | 100% | 100% |
|  | Sought care at study facilities | 100% | 100% | 100% | 100% |
|  | Sought care | 99% | 95% | 96% | 99% |
|  | Total | 97% | 94% | 95% | 93% |
| Average cases, high care seeking | Enrolled | 100% | 100% | 100% | 100% |
|  | Sought care at study facilities | 100% | 100% | 100% | 100% |
|  | Sought care | 99% | 96% | 96% | 98% |
|  | Total | 99% | 95% | 94% | 94% |
| Average cases, low care seeking | Enrolled | 100% | 100% | 100% | 100% |
|  | Sought care at study facilities | 100% | 100% | 100% | 100% |
|  | Sought care | 98% | 95% | 95% | 99% |
|  | Total | 93% | 96% | 96% | 92% |
| High cases, average care seeking | Enrolled | 100% | 100% | 100% | 100% |
|  | Sought care at study facilities | 100% | 100% | 100% | 100% |
|  | Sought care | 99% | 96% | 96% | 99% |
|  | Total | 97% | 95% | 95% | 95% |
| Low cases, average care seeking | Enrolled | 100% | 100% | 100% | 100% |
|  | Sought care at study facilities | 100% | 100% | 100% | 100% |
|  | Sought care | 99% | 97% | 97% | 97% |
|  | Total | 98% | 95% | 95% | 87% |

In the average scenario, M-estimation confidence intervals were the least precise across incidence adjustment levels (Table 4). Bootstrap percentile and Wald-type confidence intervals had very similar precision, and these were also very similar to Monte Carlo simulation intervals for incidence of enrolled cases and cases who sought care at study facilities (Table 4). Bootstrap was more precise for incidence of cases who sought care, while Monte Carlo was more precise for total incidence (Table 4). This was generally consistent across scenarios except for the low care seeking scenario, where bootstrap confidence intervals for total incidence were very wide for some simulations, resulting in lower precision on average compared to both M-estimation and Monte Carlo simulations (Figure 1, Table 4).

**Table 4.** Median and interquartile range of confidence interval width across simulation scenarios, for different incidence adjustment levels and uncertainty estimation methods.

| Scenario | Incidence adjustment level | M-estimation (median, IQR) | Bootstrap percentile (median, IQR) | Bootstrap Wald (median, IQR) | Monte Carlo (median, IQR) |
| --- | --- | --- | --- | --- | --- |
| Average cases, average care seeking | Enrolled | 0.36 (IQR: 0.35-0.38) | 0.21 (IQR: 0.20-0.22) | 0.21 (IQR: 0.21-0.22) | 0.23 (IQR: 0.22-0.24) |
|  | Sought care at study facilities | 0.52 (IQR: 0.50-0.54) | 0.30 (IQR: 0.29-0.32) | 0.31 (IQR: 0.29-0.32) | 0.28 (IQR: 0.26-0.29) |
|  | Sought care | 1.76 (IQR: 1.63-1.89) | 1.40 (IQR: 1.29-1.53) | 1.40 (IQR: 1.29-1.53) | 1.67 (IQR: 1.56-1.81) |
|  | Total | 5.65 (IQR: 5.20-6.15) | 5.13 (IQR: 4.69-5.73) | 5.06 (IQR: 4.63-5.61) | 4.69 (IQR: 4.29-5.13) |
| Average cases, high care seeking | Enrolled | 0.57 (IQR: 0.55-0.59) | 0.29 (IQR: 0.28-0.31) | 0.30 (IQR: 0.28-0.31) | 0.30 (IQR: 0.29-0.31) |
|  | Sought care at study facilities | 0.82 (IQR: 0.79-0.85) | 0.42 (IQR: 0.40-0.44) | 0.42 (IQR: 0.41-0.44) | 0.37 (IQR: 0.36-0.38) |
|  | Sought care | 2.59 (IQR: 2.43-2.76) | 1.90 (IQR: 1.76-2.06) | 1.90 (IQR: 1.76-2.06) | 2.07 (IQR: 1.94-2.24) |
|  | Total | 4.50 (IQR: 4.22-4.80) | 3.63 (IQR: 3.33-3.95) | 3.61 (IQR: 3.33-3.92) | 3.37 (IQR: 3.14-3.64) |
| Average cases, low care seeking | Enrolled | 0.25 (IQR: 0.24-0.27) | 0.16 (IQR: 0.16-0.17) | 0.17 (IQR: 0.16-0.17) | 0.18 (IQR: 0.17-0.19) |
|  | Sought care at study facilities | 0.36 (IQR: 0.35-0.38) | 0.23 (IQR: 0.23-0.24) | 0.24 (IQR: 0.23-0.25) | 0.22 (IQR: 0.21-0.22) |
|  | Sought care | 1.35 (IQR: 1.24-1.48) | 1.16 (IQR: 1.05-1.29) | 1.16 (IQR: 1.05-1.29) | 1.49 (IQR: 1.36-1.62) |
|  | Total | 6.97 (IQR: 6.21-7.76) | 7.88 (IQR: 6.74-9.28) | 7.73 (IQR: 6.53-9.95) | 6.42 (IQR: 5.70-7.12) |
| High cases, average care seeking | Enrolled | 0.92 (IQR: 0.89-0.96) | 0.43 (IQR: 0.41-0.45) | 0.43 (IQR: 0.41-0.45) | 0.41 (IQR: 0.40-0.43) |
|  | Sought care at study facilities | 1.32 (IQR: 1.27-1.37) | 0.61 (IQR: 0.58-0.64) | 0.61 (IQR: 0.59-0.64) | 0.51 (IQR: 0.49-0.53) |
|  | Sought care | 4.81 (IQR: 4.47-5.19) | 3.85 (IQR: 3.53-4.23) | 3.85 (IQR: 3.53-4.22) | 4.90 (IQR: 4.56-5.31) |
|  | Total | 15.77 (IQR: 14.47-17.18) | 14.54 (IQR: 13.20-16.04) | 14.36 (IQR: 13.00-15.79) | 13.93 (IQR: 12.77-15.18) |
| Low cases, average care seeking | Enrolled | 0.12 (IQR: 0.12-0.13) | 0.09 (IQR: 0.09-0.10) | 0.10 (IQR: 0.09-0.10) | 0.11 (IQR: 0.10-0.11) |
|  | Sought care at study facilities | 0.18 (IQR: 0.17-0.19) | 0.14 (IQR: 0.13-0.14) | 0.14 (IQR: 0.13-0.14) | 0.13 (IQR: 0.12-0.13) |
|  | Sought care | 0.52 (IQR: 0.49-0.56) | 0.43 (IQR: 0.40-0.47) | 0.43 (IQR: 0.41-0.47) | 0.42 (IQR: 0.39-0.45) |
|  | Total | 1.62 (IQR: 1.50-1.77) | 1.47 (IQR: 1.35-1.63) | 1.46 (IQR: 1.34-1.61) | 1.10 (IQR: 1.02-1.22) |

### 3.2 EFGH Results

#### 3.2.1 Associations with care seeking and Shigella etiology

In a multivariable regression model of the EFGH HUS used to parametrize the average care seeking scenarios of the simulation, we found number of days with fever (odds ratio [OR] = 1.23; 95% CI: 1.19, 1.28), number of days with vomiting (OR = 1.31; 95% CI: 1.24, 1.39), and number of loose stools on worst day of diarrhea (OR = 1.13; 95% CI: 1.09, 1.17) were all associated with increased healthcare seeking after adjusting for age and wealth quintile (Supplementary Table 2). In that same model, wealth quintile was also associated with healthcare seeking, with wealthier quintiles being more likely to seek care than poorer wealth quintiles (Supplementary Table 2). On the other hand, age (OR = 0.99; 95% CI: 0.99, 1.00) and blood in stool (OR = 1.27; 95% CI: 1.00, 1.60) did not show evidence of an association with healthcare seeking after adjusting for other variables (Supplementary Table 2). These results were consistent in the models restricted to high and low care seeking scenarios, respectively (Supplementary Table 2).

In a multivariable regression model of EFGH cases where the outcome was diarrhea attributed to *Shigella*, adjusted for all the same variables as the HUS model, we found that a higher age in months (OR = 1.06; 95% CI: 1.06, 1.07), greater number of days with fever (OR = 1.06; 95% CI: 1.03, 1.10), greater number of loose stools on worst day of diarrhea (OR = 1.08; 95% CI: 1.06, 1.10), and blood in stool (OR = 3.99; 95% CI: 3.45, 4.61) were all significantly associated with *Shigella* diarrhea (Supplementary Table 2). Number of days with vomiting was negatively associated with *Shigella* diarrhea (OR = 0.86; 95% CI: 0.81, 0.90) and wealth quintile was not significantly associated with *Shigella* diarrhea (Supplementary Table 2).

#### 3.2.2. Adjusted incidence and uncertainty estimation

Based on our simulation results, we adjusted for healthcare seeking in the EFGH data using propensity score weights truncated at the 95^th^ percentile. There was important variability in the incidence of *Shigella* diarrhea cases enrolled in EFGH across country sites, and variability in healthcare seeking behaviors are evidenced by the difference between incidence of cases who sought care at study facilities and total incidence (Figure 2). Healthcare seeking for diarrhea ranged from 16% to 55% across sites, and among those who sought care, seeking care at an EFGH facility ranged from 16% to 88% (Supplementary Table 3). Across sites, the healthcare seeking adjustment resulted in a greater percent increase in incidence for all *Shigella* diarrhea (66-766%) than for moderate-to-severe *Shigella* diarrhea (40-476%, Supplementary Figure 1).

**Figure 2.**
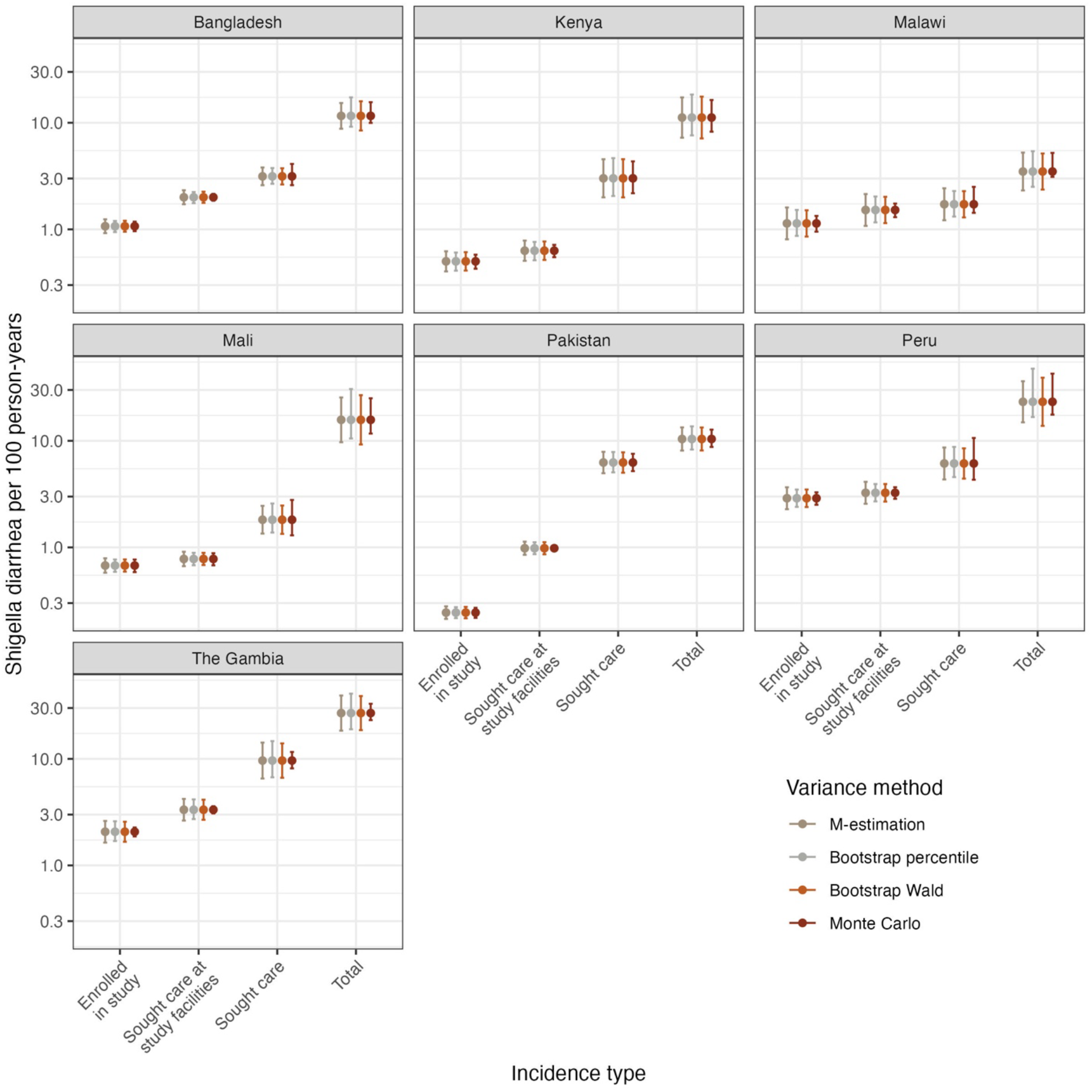
Incidence of Shigella diarrhea per 100 person-years among children 6-35 months of age in country sites included in the EFGH study, with uncertainty estimated using M-estimation, bootstrap, and Monte Carlo simulations. Note y-axis is on the log scale.

Consistent with the simulation results, M-estimation confidence intervals were generally slightly less precise than other methods for incidence for cases enrolled, cases who sought care at study facilities, and cases who sought care (Figure 2; Supplementary Table 4). However, M-estimation was slightly more precise than bootstrap for total incidence, with Wald-style confidence intervals being more precise than percentile confidence intervals (Figure 2; Supplementary Table 4). Precision of Monte Carlo simulation intervals was highly variable across sites and incidence adjustment levels; it was generally more precise than other methods, but for certain sites and incidence estimates it was the least (or close to the least) precise (Figure 2; Supplementary Table 4).

Computation times varied widely across methods, with the fastest being Monte Carlo simulations (10,000 simulations completed in 27 seconds), followed by bootstrap (1,000 resamples completed in 36 seconds), and M-estimation using *geex* being significantly slower (completed in one hour and 34 minutes).

## 4. Discussion

In this analysis, we sought to improve the healthcare seeking adjustment and the uncertainty estimation methods used in hybrid surveillance studies of infectious disease incidence. For the healthcare seeking adjustment, we found that weighting cases with the inverse of the marginal probability of healthcare seeking, as was done in prior studies of diarrheal disease (GEMS^28^ and VIDA^12^), is likely to lead to bias, and the direction of this bias is difficult to predict. Using a logistic regression model to generate propensity for healthcare seeking scores based on a set of sociodemographic and disease severity characteristics, and weighting cases using the inverse of these individual-level scores, greatly reduces bias in the estimates. However, propensity score weights tended to overestimate incidence in most scenarios, likely due to extreme weights resulting from limited data in certain strata of the covariates included in the model. This was resolved by truncating weights at the 95^th^ percentile, which resulted in approximately unbiased incidence estimates across almost all scenarios. When applied to the EFGH data, we found the healthcare seeking adjustment to result in a smaller but still important increase in incidence for moderate-to-severe *Shigella* diarrhea, which is more likely to be the primary endpoint in a vaccine trial.

To adjust for healthcare seeking, one recent typhoid fever hybrid surveillance study considered the influence of wealth and education,^8^ while two others only stratified by age and site.^9,10^ Our results suggest that, although these stratifications are likely an improvement over weighting based on the marginal probability of healthcare seeking, this may be insufficient and may have yielded biased results. After adjusting for symptoms, age, and wealth in EFGH, we found that most symptoms—but not age—significantly predicted care seeking, highlighting the importance of accounting for factors other than age. Furthermore, we found that all symptoms associated with care seeking were also associated with *Shigella* attribution among children with diarrhea. When individuals with the target pathogen (e.g., typhoid, *Shigella*) are more or less likely to seek care than other individuals with the relevant syndrome (e.g., febrile illness, diarrhea), the estimated incidence may be further biased.^13^ The propensity score approach appears to account for bias due to both general heterogeneity in care seeking as well as differential care seeking among those with the etiology of interest, as both were incorporated in our simulation.

In our simulation, we also compared how well three uncertainty estimation methods for hybrid surveillance studies captured the target of inference, which was the simulation-specific true incidence. We found that M-estimation and bootstrap confidence intervals both had at least nominal and often conservative coverage of the true total incidence across scenarios. Our findings are consistent with other simulation studies of fusion designs using M-estimation, which have found nominal coverage of confidence intervals across several estimators.^18–20^ Monte Carlo intervals had nominal coverage in certain scenarios, but in the low case scenario, for example, had substantially less than nominal coverage. All three uncertainty estimation methods produced conservative confidence intervals for incidence of enrolled cases and incidence of cases who sought care at study facilities, both of which we estimated directly in the population we were trying to make an inference in (e.g., enrolled cases). This is in contrast to the incidence of cases who sought care and total incidence, which we estimated based on a sample from a super population. When applied to the EFGH data, we again found that Monte Carlo simulation intervals were often most precise, and that bootstrap and M-estimation confidence intervals were similar.

We also evaluated each uncertainty estimation method’s efficiency and ease of implementation. When implemented using the *geex* package.^27^ M-estimation was significantly more computationally intensive than the other methods, taking more than an hour to complete compared to less than a minute for Monte Carlo and bootstrap. Additionally, M-estimation implementation is not trivial. Though simplified by the functions in the *geex* package, it requires specific data formatting and an understanding of the structure of the estimating functions, which can be difficult to specify depending on the complexity of the analysis at hand. Although bespoke M-estimation functions or implementation through the *delicatessen* package in Python^29^ may be more computationally efficient than *geex*, this would require specialized knowledge and may not be feasible for some analysts. Bootstrap implementation is comparatively straightforward and thus may be a preferred approach.

Uncertainty estimation across prior hybrid surveillance studies has been inconsistent. The initial GEMS analysis estimated uncertainty around incidence using the delta method, which appropriately accounts for uncertainty when estimating several unknown quantities from different data sets, and is mathematically equivalent to M-estimation.^28^ A re-analysis of GEMS, as well as VIDA, instead used bootstrapping to first estimate uncertainty around the attributable fraction for each pathogen, and then used the bootstrapped estimates, the estimate of all cause diarrhea incidence, and the delta method to estimate uncertainty around etiology-specific incidence.^2,12^ Two of the aforementioned typhoid studies used Monte Carlo simulations to sample from a distribution of each adjustment probability and from a distribution of cases,^8,9^ while the third used a Bayesian mixture model.^10^

An important limitation of this analysis is the potential recall bias in the HUS data, from which the healthcare seeking adjustments are estimated. Caregivers who brought their child to care are probably more likely to recall their symptoms, or to recall them as more severe, than those who did not bring their child to care. Additionally, symptoms reported by facility cases are limited to those that occurred in the lead up to the decision to seek care. On the other hand, symptoms reported in the HUS, which covered a two-week recall period, may include symptoms that occurred for only part of the episode (if ongoing at the time of the survey) or for the entire episode (if resolved) and therefore not necessarily limited to before caregivers decided whether to seek care. This may impact the accuracy of the healthcare seeking weights generated by the propensity score model, which rely on the data reported by caregivers. In our simulation we implicitly assume no bias in the HUS responses and that the propensity score model for healthcare seeking is correctly specified. These assumptions are likely not realistic and their impact on estimated incidence rates should be evaluated in future work. Similarly, duration of symptoms may be an important determinant of care seeking, but this was not incorporated in our model given that this is unknown for HUS respondents for whom symptoms have not yet resolved, and for all facility cases. Future work should evaluate deriving propensity for care seeking weights from a time-to-event model incorporating the probability of seeking care each day after the start of symptoms.

In conclusion, we found that propensity for healthcare seeking scores, with truncating of extreme weights, can be used to appropriately adjust for healthcare seeking behaviors in hybrid studies; these are a great improvement over adjusting using the marginal or stratified probability for healthcare seeking which is likely to yield biased estimates. Additionally, M-estimation or bootstrap may be an appealing option for uncertainty estimation in hybrid studies as they both provide nominal coverage and reasonable precision. However, bootstrap confidence intervals may be a preferable approach considering M-estimation’s computation and implementation challenges. We found that Monte Carlo did not consistently provide appropriate coverage, and it requires unverifiable assumptions of the distribution of adjustment parameters.

Our results illustrate an approach to accurately estimate infectious disease incidence from hybrid surveillance studies with appropriate uncertainty, removing the need to conduct resource intensive cohort studies. The incidence estimates generated with these methods for the EFGH study will be crucial to informing upcoming *Shigella* vaccine trials,^22^ and the approach should be widely applicable to future hybrid studies of other infectious diseases.

## 5. Acknowledgements

The authors thank the entire EFGH Study team, including the coordination teams and the EFGH consortium, the children who participated in these studies and their families, and the dedicated physicians, nurses, scientists, and staff at each study site.

## 7. Data availability

The corresponding R code for the simulation study and for analysis of the EFGH data can be found on GitHub (https://github.com/mariagq922/hybrid-incidence). The EFGH statistical analysis plan (https://clinicaltrials.gov/study/NCT06047821) and study protocol (https://academic.oup.com/ofid/issue/11/Supplement_1) were made publicly available, and the EFGH dataset is deidentified and anonymized and can be found in Vilvi from December 2025 at https://doi.org/10.25934/PR00011860.

