## Supplementary Materials for "Comparing existing and novel methods for estimating etiology-specific diarrheal disease incidence in hybrid surveillance studies"

### Appendix 1: Illustration of advantage of propensity score model

Supplementary Table A1 shows a simplified example with dummy data to illustrate how the propensity score model works. In this example, suppose we find in the healthcare utilization survey (HUS) that 5% of cases with mild diarrhea access care, 25% of cases with moderate diarrhea access care, and 90% of cases with severe diarrhea access care. Then, suppose we enroll 100 cases in one of the study facilities, out of which 10 have mild diarrhea, 30 have moderate diarrhea, and 60 have severe diarrhea. We can then use the propensities to seek care for each disease severity strata to upweight for the proportion with the same disease severity that did not access care. Then, we can sum across the adjusted disease severity strata to find that the 'true' number of cases is 387 (200+120+67). However, if we had estimated a single propensity to seek care across all severity strata, we would have estimated a falsely inflated number of cases of 958 (100*(1/0.10)).

*Supplementary Table A1. Simplified illustration with dummy data of logic behind propensity score model.*

| ***Disease severity*** | **A. Number in HUS** | **B. Number in HUS who accessed care** | **C. Propensity to seek care (B/A)** | **D. Weight (1/C)** | **E. Facility cases enrolled** | **Adjusted number of cases (E x D)** |
|---:|---:|---:|---:|---:|---:|---:|
| *Mild* | 1000 | 50 | 0.05 | 20.0 | 10 | 200 |
| *Moderate* | 100 | 25 | 0.25 | 4.0 | 30 | 120 |
| *Severe* | 50 | 45 | 0.90 | 1.1 | 60 | 67 |
| ***All diarrhea*** | 1150 | 120 | 0.10 | 9.6 | 100 | 958 |

The model's advantage over a simple stratification is that we can estimate propensity scores (i.e., propensity to seek care) based on many variables. The model used in EFGH incorporates age, sex, country site, wealth quintile, number of days with fever, number of days with vomit, number of times vomited on worst day, number of loose stools on worst day, and reported blood in stool. These variables were selected due to their hypothesized association with healthcare seeking behaviors and because they were measured similarly in the enrollment and healthcare utilization surveys (e.g., caregiver report).

### Appendix 2: Variable selection for EFGH propensity score model

To maximize predictive accuracy of the model, all variables with a hypothesized association with healthcare seeking behaviors that were captured by both the facility enrollment survey and the HUS were considered for inclusion as predictors. Each variable was then evaluated to assess whether it may have been ascertained differently between surveys. Specifically, we only included symptom variables that were measured by caregiver report in both settings, as we expect a clinician assessment (only available among facility cases) to differ significantly from that of a caregiver. Furthermore, we qualitatively compared the distribution of potentially eligible variables between facility cases and children in the HUS that reported accessing care. Variables with meaningfully different distributions were assumed to have been captured differently and were excluded. All variables considered and their reason for exclusion, if any, are described in Supplementary Table A2.

To select which of all possible two-way interactions to include in the model, we used cross-validated LASSO to identify two-way interactions that maximized the predictive accuracy of the model. Since two categorical variables were included (country and wealth quintile), we used group-LASSO, which allowed us to require that interactions with each level of a categorical variable either all be selected, or all be excluded. We also leveraged this functionality to group all main terms in the model such that these were always all selected, and LASSO only selected from among the two-way interactions. We used 10-fold cross-validation to estimate the lambda (i.e., regularization parameter) that yielded the minimum cross-validation error, which determined the final form of the model, shown below. This analysis was done using R package *gglasso*.[1]

*Supplementary Table A2. Demographic and symptom variables included in both HUS and facility case enrollment survey, whether each was included in the propensity score model, and reason for exclusion.*

| VARIABLE | INCLUDED (YES/NO) | REASON FOR EXCLUSION |
|---|---|---|
| Age (months) | Yes | |
| Sex | Yes | |
| Country site | Yes | |
| Wealth index | No | Captured by wealth quintile, resulted in some positivity issues (extreme weights) |
| Wealth quintile | Yes | |
| Any fever | No | Captured by number of days with fever |
| Number of days with fever | Yes | |
| Any vomiting | No | Captured by number of days with vomiting |
| Number of days with vomiting | Yes | |
| Number of times vomited on worst day of vomiting | Yes | |

| | | |
|---|---|---|
| Number of loose stools on worst day of diarrhea | **Yes** | |
| Blood in stool | **Yes** | |
| General condition (Lethargic or Unconscious, Restless/Irritable, Well/Alert) | **No** | The distribution of responses among those who sought care in HUS vs enrolled cases is very different, suggesting responses for cases were informed by clinician assessment, which is not representative of caregiver assessment in HUS. |
| Ability to drink (Drinks poorly/Unable to drink, Drinks eagerly/Thirsty, Normal/Not thirsty) | **No** | The distribution of responses among those who sought care in HUS vs enrolled cases is very different, suggesting responses for cases were informed by clinician assessment, which is not representative of caregiver assessment in HUS. |
| Sunken eyes | **No** | Too subjective. Clinician assessment not equivalent to caregiver assessment. |
| Wrinkled skin | **No** | Too subjective. Clinician assessment not equivalent to caregiver assessment. |
| Number of children aged <5 years in the household | **No** | For Mali, distribution of responses between HUS and case data was very different, suggesting the question is likely being captured differently in both surveys (complicated by polygamist/multi-family compounds). |

```
model <- hus_careseek_mad ~ countryKenya + countryMalawi + countryMali + countryPakistan + countryPeru +
countryTheGambia +
  age +
  sex +
  fever_days +
  vomit_days +
  vomit_times +
  stools +
  blood +
  wealth_quintile2 + wealth_quintile3 + wealth_quintile4 + wealth_quintile5 +
  (countryKenya + countryMalawi + countryMali + countryPakistan + countryPeru + countryTheGambia)*age +
  (countryKenya + countryMalawi + countryMali + countryPakistan + countryPeru + countryTheGambia)*fever_days +
  (countryKenya + countryMalawi + countryMali + countryPakistan + countryPeru + countryTheGambia)*vomit_days +
  (countryKenya + countryMalawi + countryMali + countryPakistan + countryPeru + countryTheGambia)*vomit_times +
  (countryKenya + countryMalawi + countryMali + countryPakistan + countryPeru + countryTheGambia)*stools +
  age*sex +
  age*fever_days +
  age*vomit_days +
  age*vomit_times +
  age*stools +
  age*(wealth_quintile2 + wealth_quintile3 + wealth_quintile4 + wealth_quintile5) +
  fever_days*vomit_days +
  fever_days*vomit_times +
  fever_days*stools +
  fever_days*blood +
  fever_days*(wealth_quintile2 + wealth_quintile3 + wealth_quintile4 + wealth_quintile5) +
  vomit_days*vomit_times +
  vomit_times*blood +
  stools*blood +
  stools*(wealth_quintile2 + wealth_quintile3 + wealth_quintile4 + wealth_quintile5)
```

### Appendix 3: Supplementary Tables and Figures

*Supplementary Table 1. Parameters for simulated hybrid studies, based on EFGH data.*

| Parameter | Value | Distribution |
|---|---|---|
| **For population at risk** | | |
| **Number of catchment area clusters** | 500 | Fixed |
| **Proportion of clusters enumerated** | 0.6 | Binomial |
| **Households per cluster** | 100-200 | Uniform |
| **Proportion of HH enumerated** | 0.97 | Binomial |
| **Proportion of HH with 1+ kid <5y** | 0.4 | Binomial |
| **Avg number of kids <5y in HH with 1+ kid <5y** | 1.4 | Poisson |
| **Proportion of kids 6-35m reporting diarrhea** | 0.1 | Binomial |
| **Healthcare seeking among those with diarrhea** | Propensity score based on regression models in Supplementary Table 2 | Binomial |
| **Cluster-level variability in healthcare seeking** | 0.1 | Normal[2] |
| **Proportion seeking care at study facilities** | 0.4 | Binomial |
| **For diarrhea cases** | | |
| **Number of study facilities** | 10 | Fixed |
| **Proportion of cases enrolled in study** | 0.7 | Binomial |
| ***Shigella* etiology** | Propensity score based on regression model in Supplementary Table 2 | Binomial |
| **For both (pop. at risk & cases)** | | |
| **Avg stools (by age group)[1]** | 4.8, 4.6, 4.5, 4.4, 4.5 | Poisson |
| **Probability of blood in stool (by age group)[1]** | 0.05, 0.07, 0.08, 0.10, 0.09 | Binomial |
| **Avg days with fever (by age group)[1]** | 1.5, 1.3, 1.3, 1.3, 1.2 | Poisson |
| **Avg days with vomiting (by age group)[1]** | 0.8, 0.7, 0.5, 0.5, 0.5 | Poisson |
| **Individual-level variability in healthcare seeking** | 0.1 | Normal[2] |

[1]Age groups: 6-11, 12-17, 18-23, 24-29, and 30-35 months

[2]Value is standard deviation applied to a normal distribution with mean 0

*Supplementary Table 2. Odds ratios from multivariable regression models use to parametrize simulated hybrid studies.*

| | **Outcome: Seeking care**[1] | | | **Outcome: *Shigella* etiology**[2] |
|---|---|---|---|---|
| **Covariates** | Average across EFGH sites (OR, 95% CI) | High care seeking site (OR, 95% CI) | Low care seeking site (OR, 95% CI) | (OR, 95% CI) |
| **(Intercept)** | 0.20 (0.15, 0.27) | 0.21 (0.11, 0.39) | 0.02 (0.00, 0.17) | 0.05 (0.04, 0.06) |
| **Age in months** | 0.99 (0.99, 1.00) | 0.99 (0.98, 1.01) | 0.99 (0.95, 1.02) | 1.06 (1.06, 1.07) |
| **Number of days with fever** | 1.23 (1.19, 1.28) | 1.59 (1.43, 1.76) | 1.29 (1.07, 1.55) | 1.06 (1.03, 1.10) |
| **Number of days with vomiting** | 1.31 (1.24, 1.39) | 1.27 (1.10, 1.47) | 1.46 (1.15, 1.85) | 0.86 (0.81, 0.90) |
| **Number of loose stools on worst day of diarrhea** | 1.13 (1.09, 1.17) | 1.31 (1.19, 1.45) | 1.03 (0.81, 1.30) | 1.08 (1.06, 1.10) |
| **Blood in stool** | 1.27 (1.00, 1.60) | 2.08 (0.94, 4.95) | 2.53 (0.73, 7.84) | 3.99 (3.45, 4.61) |
| **Wealth quintile** | | | | |
| **1 (poorest)** | Ref. | Ref. | Ref. | Ref. |
| **2** | 1.01 (0.82, 1.23) | 0.88 (0.58, 1.32) | 3.21 (0.62, 59.0) | 0.98 (0.83, 1.15) |
| **3** | 1.19 (0.97, 1.46) | 1.18 (0.79, 1.76) | 4.80 (0.92, 88.4) | 0.97 (0.82, 1.14) |
| **4** | 1.52 (1.24, 1.86) | 1.35 (0.92, 1.99) | 8.57 (1.63, 158) | 0.92 (0.76, 1.10) |
| **5 (wealthiest)** | 0.95 (0.76, 1.19) | 1.52 (0.98, 2.37) | 21.3 (1.55, 567) | 0.99 (0.80, 1.23) |

OR = odds ratio, CI = confidence interval

[1]Estimated on EFGH HUS data.

[2]Estimated on EFGH facility case data.

*Supplementary Table 3. Sociodemographic and clinical characteristics of EFGH healthcare utilization survey respondents (children 6-35 months old who experienced diarrhea in the past two weeks).*

| | | | **Sought care (row %)** | | | **Among those who sought care, sought care at EFGH facility (row %)** | |
|---|---|---|---|---|---|---|---|
| **Variable** | **N = 4,794 (col %)** | **N** | **No** N = 3,072 | **Yes** N = 1,722 | **N** | **No** N = 1,139 | **Yes** N = 583 |
| **Country** | | 4,794 | | | 1,731 | | |
| Bangladesh | 816 (17%) | | 640 (78%) | 176 (22%) | | 64 (36%) | 112 (64%) |
| Kenya | 574 (12%) | | 402 (70%) | 172 (30%) | | 136 (79%) | 36 (21%) |
| Malawi | 280 (5.8%) | | 142 (51%) | 138 (49%) | | 16 (12%) | 122 (88%) |
| Mali | 528 (11%) | | 444 (84%) | 84 (16%) | | 48 (57%) | 36 (43%) |
| Pakistan | 1,283 (27%) | | 576 (45%) | 707 (55%) | | 596 (84%) | 111 (16%) |
| Peru | 235 (4.9%) | | 186 (79%) | 49 (21%) | | 23 (47%) | 26 (53%) |
| The Gambia | 1,078 (22%) | | 673 (62%) | 405 (38%) | | 265 (65%) | 140 (35%) |
| **Wealth quintile** | | 4,794 | | | 1,731 | | |
| 1 (poorest) | 798 (17%) | | 525 (66%) | 273 (34%) | | 184 (67%) | 89 (33%) |
| 2 | 1,140 (24%) | | 752 (66%) | 388 (34%) | | 245 (63%) | 143 (37%) |
| 3 | 1,045 (22%) | | 660 (63%) | 385 (37%) | | 266 (69%) | 119 (31%) |
| 4 | 978 (20%) | | 567 (58%) | 411 (42%) | | 287 (70%) | 124 (30%) |
| 5 (wealthiest) | 833 (17%) | | 559 (67%) | 274 (33%) | | 166 (61%) | 108 (39%) |
| **Age in months[1]** | 18 (12, 25) | 4,794 | 18 (12, 25) | 17 (12, 24) | 1,731 | 17 (12, 25) | 16 (11, 23) |
| **Any fever** | | 4,788 | | | 1,728 | | |
| No | 2,265 (47%) | | 1,685 (74%) | 580 (26%) | | 377 (65%) | 203 (35%) |
| Yes | 2,523 (53%) | | 1,375 (54%) | 1,148 (46%) | | 769 (67%) | 379 (33%) |
| **Any vomit** | | 4,794 | | | 1,731 | | |
| No | 3,326 (69%) | | 2,349 (71%) | 977 (29%) | | 670 (69%) | 307 (31%) |
| Yes | 1,468 (31%) | | 714 (49%) | 754 (51%) | | 478 (63%) | 276 (37%) |
| **Number of loose stools on worst day of diarrhea†** | 4 (4, 5) | 4,776 | 4 (3, 5) | 4 (4, 5) | 1,723 | 4 (4, 5) | 5 (4, 6) |
| **Blood in stool** | | 4,787 | | | 1,730 | | |
| No | 4,440 (93%) | | 2,869 (65%) | 1,571 (35%) | | 1,061 (68%) | 510 (32%) |
| Yes | 347 (7.2%) | | 188 (54%) | 159 (46%) | | 87 (55%) | 72 (45%) |

†Median (IQR)

*Supplementary Table 4. EFGH incidence estimates and confidence interval widths using different uncertainty estimation methods.*

| | | | *95% Confidence or simulation interval width* | | | |
|---|---|---|---|---|---|---|
| *Incidence adjustment level* | *Country* | *Incidence per 100 person-years* | *M-estimation* | *Bootstrap percentile* | *Bootstrap Wald* | *Monte Carlo* |
| Enrolled in study | Bangladesh | 1.07 | 0.32 | 0.26 | 0.26 | 0.22 |
| | Kenya | 0.50 | 0.22 | 0.20 | 0.20 | 0.15 |
| | Malawi | 1.14 | 0.80 | 0.65 | 0.65 | 0.39 |
| | Mali | 0.68 | 0.21 | 0.18 | 0.18 | 0.18 |
| | Pakistan | 0.24 | 0.07 | 0.06 | 0.06 | 0.05 |
| | Peru | 2.89 | 1.38 | 1.09 | 1.10 | 0.77 |
| | The Gambia | 2.07 | 0.98 | 0.90 | 0.90 | 0.40 |
| Sought care at study facilities | Bangladesh | 2.00 | 0.61 | 0.48 | 0.48 | 0.29 |
| | Kenya | 0.63 | 0.28 | 0.25 | 0.25 | 0.17 |
| | Malawi | 1.52 | 1.06 | 0.87 | 0.88 | 0.44 |
| | Mali | 0.78 | 0.24 | 0.21 | 0.21 | 0.20 |
| | Pakistan | 0.98 | 0.28 | 0.25 | 0.26 | 0.11 |
| | Peru | 3.25 | 1.55 | 1.23 | 1.24 | 0.81 |
| | The Gambia | 3.33 | 1.57 | 1.44 | 1.46 | 0.52 |
| Sought care | Bangladesh | 3.14 | 1.21 | 1.08 | 1.11 | 1.51 |
| | Kenya | 3.01 | 2.55 | 2.60 | 2.56 | 2.16 |
| | Malawi | 1.72 | 1.21 | 0.97 | 0.99 | 1.07 |
| | Mali | 1.82 | 1.10 | 1.20 | 1.13 | 1.49 |
| | Pakistan | 6.26 | 2.92 | 2.79 | 2.77 | 2.35 |
| | Peru | 6.13 | 4.31 | 4.20 | 4.08 | 6.32 |
| | The Gambia | 9.64 | 7.67 | 7.96 | 7.31 | 3.48 |
| Total | Bangladesh | 11.60 | 6.53 | 8.12 | 7.44 | 5.63 |
| | Kenya | 11.19 | 10.05 | 10.77 | 10.45 | 8.10 |
| | Malawi | 3.49 | 2.93 | 2.90 | 2.79 | 2.11 |
| | Mali | 15.74 | 15.80 | 20.12 | 17.54 | 13.37 |
| | Pakistan | 10.40 | 5.27 | 5.37 | 5.22 | 3.96 |
| | Peru | 23.24 | 21.30 | 30.82 | 25.29 | 25.08 |
| | The Gambia | 26.89 | 20.87 | 21.95 | 20.22 | 9.79 |

*Supplementary Table 5. Bias in simulated observed incidence point estimates compared to the truth across scenarios, for different incidence adjustment levels.*

| *Scenario* | *Incidence adjustment level* | *True incidence (mean, range)* | *Estimated incidence (mean, range)* | *Relative difference in estimated incidence (mean, 95% CI)* |
|---|---|---|---|---|
| Average cases, average care seeking | Enrolled | 1.57 (1.30-1.95) | 1.57 (1.31-1.96) | -0.00% (95% CI: -0.06, 0.06) |
| | Sought care at study facilities | 2.24 (1.91-2.71) | 2.24 (1.88-2.71) | 0.05% (95% CI: -0.07, 0.17) |
| | Sought care | 5.59 (4.82-6.65) | 5.61 (4.53-7.27) | 0.25% (95% CI: -0.14, 0.64) |
| | Total | 15.78 (13.62-18.15) | 15.89 (12.05-22.79) | 0.71% (95% CI: 0.20, 1.22) |
| Average cases, high care seeking | Enrolled | 2.68 (2.26-3.19) | 2.68 (2.27-3.16) | -0.02% (95% CI: -0.07, 0.04) |
| | Sought care at study facilities | 3.83 (3.29-4.47) | 3.83 (3.26-4.48) | 0.01% (95% CI: -0.09, 0.11) |
| | Sought care | 9.55 (8.27-11.47) | 9.59 (7.67-11.73) | 0.33% (95% CI: 0.03, 0.63) |
| | Total | 15.77 (13.67-18.55) | 15.67 (12.44-20.36) | -0.65% (95% CI: -1.00, -0.30) |
| Average cases, low care seeking | Enrolled | 1.00 (0.80-1.27) | 1.00 (0.80-1.28) | -0.02% (95% CI: -0.07, 0.04) |
| | Sought care at study facilities | 1.43 (1.18-1.73) | 1.42 (1.16-1.82) | -0.02% (95% CI: -0.17, 0.13) |
| | Sought care | 3.57 (3.07-4.23) | 3.58 (2.65-4.89) | 0.30% (95% CI: -0.20, 0.80) |
| | Total | 15.77 (13.64-18.77) | 15.48 (10.70-28.27) | -1.85% (95% CI: -2.57, -1.12) |
| High cases, average care seeking | Enrolled | 4.70 (3.97-5.62) | 4.70 (3.99-5.73) | -0.00% (95% CI: -0.06, 0.06) |
| | Sought care at study facilities | 6.71 (5.75-7.95) | 6.71 (5.75-8.17) | -0.02% (95% CI: -0.10, 0.06) |
| | Sought care | 16.78 (14.59-19.42) | 16.79 (13.46-22.02) | 0.05% (95% CI: -0.31, 0.40) |
| | Total | 47.30 (40.98-54.54) | 47.57 (35.81-67.42) | 0.55% (95% CI: 0.08, 1.02) |
| Low cases, average care seeking | Enrolled | 0.36 (0.26-0.47) | 0.36 (0.26-0.46) | -0.00% (95% CI: -0.06, 0.06) |
| | Sought care at study facilities | 0.51 (0.38-0.65) | 0.51 (0.37-0.69) | -0.09% (95% CI: -0.33, 0.15) |
| | Sought care | 1.28 (1.02-1.55) | 1.28 (0.90-1.78) | -0.18% (95% CI: -0.68, 0.32) |
| | Total | 3.61 (3.02-4.26) | 3.62 (2.45-5.19) | 0.25% (95% CI: -0.38, 0.88) |

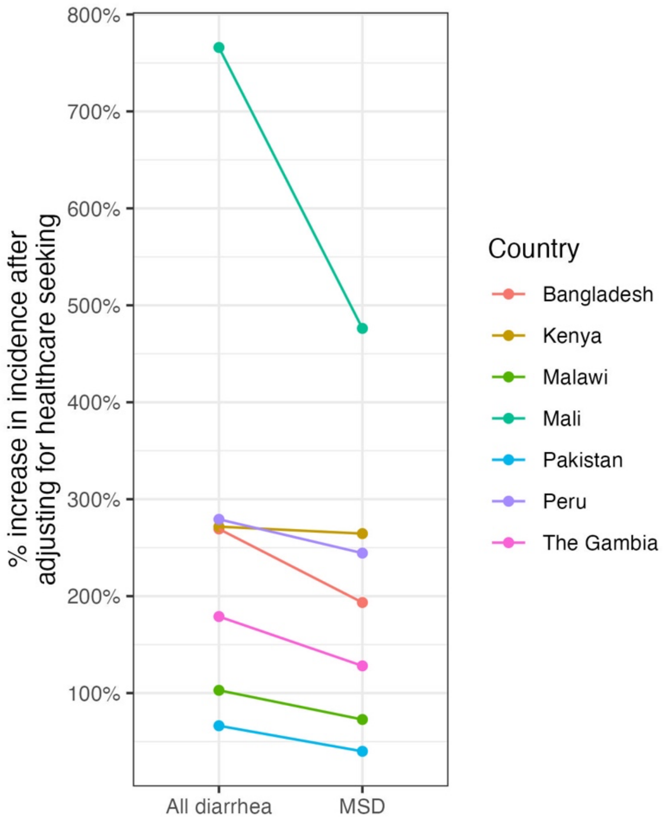

*Supplementary Figure 1. Percent increase in incidence due to healthcare seeking adjustment for all Shigella diarrhea and moderate-to-severe Shigella diarrhea (MSD) at each EFGH study site.*

Here, the comparison is between incidence of *Shigella* cases who sought care and all *Shigella* cases. Healthcare seeking adjustment was done using propensity for healthcare seeking weights truncated at the 95$^{th}$ percentile. MSD is defined as diarrhea with dysentery, dehydration, or hospitalization, in alignment with the GEMS study.[2]
